# Dynamic Macro–Microvascular Coupling During Cerebrovascular Reactivity Revealed by Simultaneous TCD and DCS

**DOI:** 10.64898/2026.11.23.26368800

**Authors:** Zhe Ji, Hui Su, Yiwen Wang, Yiqin Han, Zhe Li, Xiangyu Cao

## Abstract

**Background:** Transcranial Doppler (TCD) measures flow velocity in large intracranial arteries but provides limited information on microvascular perfusion.Diffuse correlation spectroscopy (DCS) enables continuous, noninvasive assessment of cortical microvascular blood flow. We compared macrovascular and microvascular responses during cerebrovascular reactivity testing.

**Methods:** Forty-four healthy adults (28 men; mean age, 27.0±3.8 years) underwent simultaneous bilateral TCD and DCS during a breath-holding task (1-minute baseline, 30-second breath hold, and 1-minute recovery). Middle cerebral artery flow velocity and cortical microvascular blood flow index were normalized to baseline (rCBFv and rCBFi, respectively). Temporal responses were compared using Pearson correlation and Bland–Altman analyses. Response amplitude, rise time, and recovery time were also assessed.

**Results:** TCD and DCS showed concordant response trajectories during breath holding and recovery. Correlations between normalized temporal responses were strong in both hemispheres (left,r=0.988±0.009;right, r=0.990±0.008; both P<0.001). Bland–Altman analysis showed minimal systematic bias and no proportional bias. No significant interhemispheric differences were observed in response amplitude, rise time, or recovery time. DCS-derived rCBFi showed greater response amplitudes and variability than TCD-derived rCBFv (left, 1.80±0.34 vs 1.63±0.15; right, 1.72±0.27 vs 1.63±0.14). Parameter-level analyses showed strong correlations and minimal bias for response amplitude, rise time, and recovery time.

**Conclusions:** TCD and DCS demonstrated strong temporal coupling during cerebrovascular reactivity testing, while DCS detected larger response amplitudes than TCD. These findings suggest that macrovascular velocity and cortical microvascular perfusion provide complementary information about cerebrovascular responses.

## Introduction

Cerebrovascular dysfunction is increasingly recognized as an important contributor to neurologic disorders, including ischemic stroke, cerebral small vessel disease, and vascular cognitive impairment^1,2^. Cerebrovascular reactivity (CVR), defined as the capacity of cerebral vessels to respond to vasoactive or metabolic stimuli, is a key marker of vascular reserve and cerebrovascular health. Impaired CVR has been associated with adverse clinical outcomes across a broad range of neurologic conditions^3^. Reliable CVR assessment is therefore important for characterizing cerebrovascular function, monitoring disease progression, and identifying patients at increased hemodynamic risk ^4,5^.

Transcranial Doppler ultrasonography (TCD) is widely used for CVR assessment by measuring middle cerebral artery blood flow velocity during hypercapnic challenges or breath-holding maneuvers^6^. However, TCD primarily reflects flow velocity in large intracranial arteries rather than tissue-level perfusion, limiting its ability to characterize downstream microvascular responses^7^ . Its performance may also be affected by operator dependence, insonation angle, probe stability, and acoustic window quality, which can limit reproducibility and broader use for continuous monitoring.

Diffuse correlation spectroscopy (DCS) is an emerging near-infrared optical technique that enables continuous, noninvasive monitoring of cerebral microvascular blood flow^8^. By measuring temporal fluctuations in scattered light caused by red blood cell motion, DCS provides an index of cortical microvascular perfusion and is well suited for bedside hemodynamic monitoring. Because TCD and DCS interrogate different vascular compartments, they may provide complementary information regarding macrovascular and microvascular components of CVR.

Hypercapnia induces vasodilation across the cerebrovascular tree, increasing both upstream arterial inflow and downstream microvascular perfusion^9,10^.The breath-holding test is a simple, noninvasive hypercapnic stimulus that induces transient CO₂ accumulation and is commonly used to assess CVR^11,12^. Although both middle cerebral artery flow velocity and cortical microvascular perfusion are expected to increase during breath holding, their temporal coupling and compartment-specific response characteristics have not been well defined.

In this study, we used simultaneous TCD and DCS monitoring during breath-holding stimulation to characterize dynamic cerebrovascular responses across macrovascular and microvascular compartments. This multimodal approach allowed concurrent assessment of middle cerebral artery flow velocity and cortical microvascular perfusion, enabling direct comparison of response magnitude and temporal dynamics. We hypothesized that macrovascular flow velocity and microvascular perfusion would exhibit coordinated temporal responses during CVR testing while demonstrating compartment-specific differences in response magnitude.

## Methods

### Patient Cohort

A total of 44 healthy adults aged 18–40 years were recruited from 1st Medical Center Chinese PLA General Hospital between January and March 2026. The mean age was 26.98 ± 3.80 years, with a mean height of 171.25 ± 8.00 cm and mean weight of 66.08 ± 11.96 kg; 28 participants (64%) were male. Individuals with a history of cerebrovascular disease (stroke, intracranial surgery, arterial stenosis, aneurysm, or vascular malformation) or major systemic illness were excluded. Participants with conditions that could interfere with TCD measurement at the temporal window were also excluded. All participants provided written informed consent.

### TCD

Bilateral middle cerebral artery (MCA) blood flow velocity was measured using a transcranial Doppler (TCD) system (DWL Elektronische GmbH, Sipplingen, Germany). All examinations were performed by a single experienced sonographer. The probes were secured with a head frame over the bilateral temporal acoustic windows. Before each recording, probe position and insonation angle were optimized to obtain stable MCA signals. The mean insonation depth was 53.7 ± 3.3 mm, and data were acquired at a sampling frequency of 100 Hz .

### DCS

Cerebral microvascular blood flow was quantified using diffuse correlation spectroscopy (DCS), a non-invasive optical technique based on the detection of temporal fluctuations of multiply scattered coherent near-infrared light in biological tissue. The self-developed DCS (Figure 1A) was utilized for *in vivo* measurements, which was described in our previous studies^13^. The device is briefly described here. It uses a long coherence laser (Shanghai Laser & Optics Century Co., Ltd., China) at 785 nm as irradiation source, along with a single-photon avalanche photodiode detector (Chang-Sha Lubang Photonic Technology Co., LTD, China) for detection. The software correlator in our device is designed based on a data acquisition board (PCIe-6612, National Instruments Inc., USA) and a custom software application (LabVIEW, National Instruments, USA). An optical probe was positioned on the participants’ forehead at the frontal tuber region.The correlation diffusion model was used to derive a cerebral blood flow index (CBFi), which reflects relative changes in cortical microvascular blood flow^8^.

**Figure 1.**
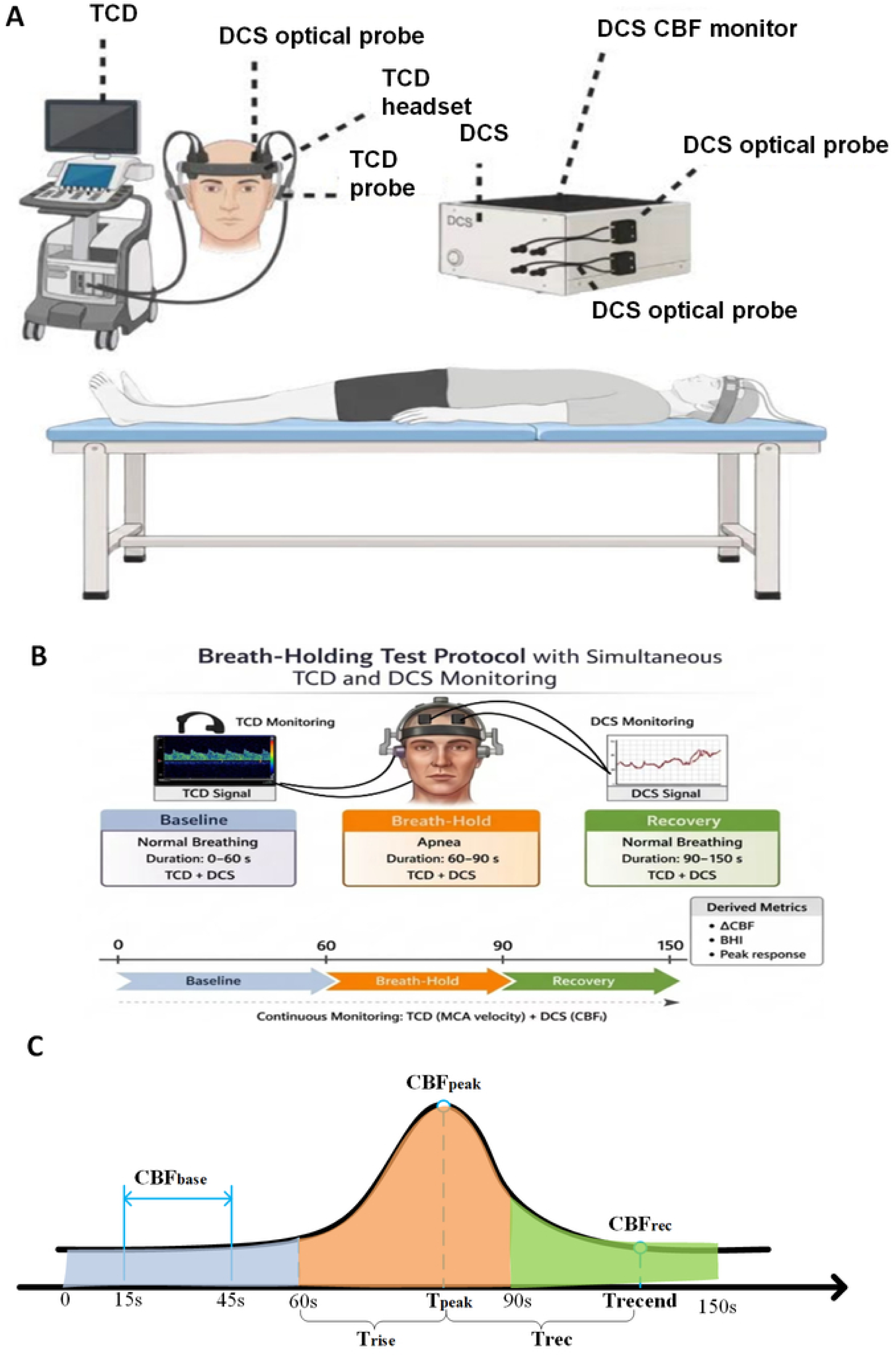
Schematic illustration of the breath-holding test (BHT) and derivation of analytical parameters. (A) Placement of transcranial Doppler (TCD) and diffuse correlation spectroscopy (DCS) probes on the human head. (B) Flowchart of the standardized physiological measurement protocol for each participant. (C) Definition of key parameters for data analysis.

### Cerebrovascular Reactivity Testing

Participants underwent a breath-holding protocol in the supine position within a quiet, temperature-controlled room maintained at 23°C (Figure 1A). Before the experiment, participants completed a baseline demographic questionnaire and received standardized instructions, including maintaining head stability to minimize motion artifacts and avoiding deep breathing during baseline recordings. After a 1-minute baseline recording, participants were instructed to perform a 30-second end-expiratory breath hold, followed by a 1-minute recovery period (Figure 1B). Trials were repeated when the breath-hold duration was less than 25 seconds or when motion artifacts significantly affected signal quality. A maximum of three attempts were allowed to obtain two valid trials. The first valid trial was used for the primary analysis.

### Dynamic time warping analysis

To quantify waveform similarity in the presence of non–uniform temporal lags between large–artery TCD-derived MCA-CBFv and microvascular DCS-derived CBFi hemodynamic responses, dynamic time warping (DTW) was adopted in this study during breath-holding-induced cerebrovascular stimulation. DTW has been previously applied to compare cerebral hemodynamic time–series in functional magnetic resonance imaging (fMRI) studies^14^. DTW is an algorithm that is employed to find an optimal alignment between two time series signals by stretching both of them in time while minimizing the distance between corresponding points in the two time series signals.

Specifically, the measured MCA–CBFV and DCS–derived CBFi signals (Figure 2A, B) were Z–score normalized to mitigate the impact of amplitude discrepancies on the resulting warping path. DTW was then implemented to identify the optimal warping path, which allows local stretching or compression along the time axis; the cumulative DTW distance derived from this path was computed to quantify waveform similarity between the two hemodynamic traces. The resulting DTW–aligned sequences are visualized in Figure 2C, D. The obtained warping path was further applied to the smoothed signals to produce time–aligned rCBFv and rCBFi sequences (Figure 2E, F), which served as input for subsequent correlation analyses. It should be noted that correlation coefficients obtained from DTW–aligned sequences represent association based on waveform matching, rather than true temporal synchrony within the original time domain.

**Figure 2.**
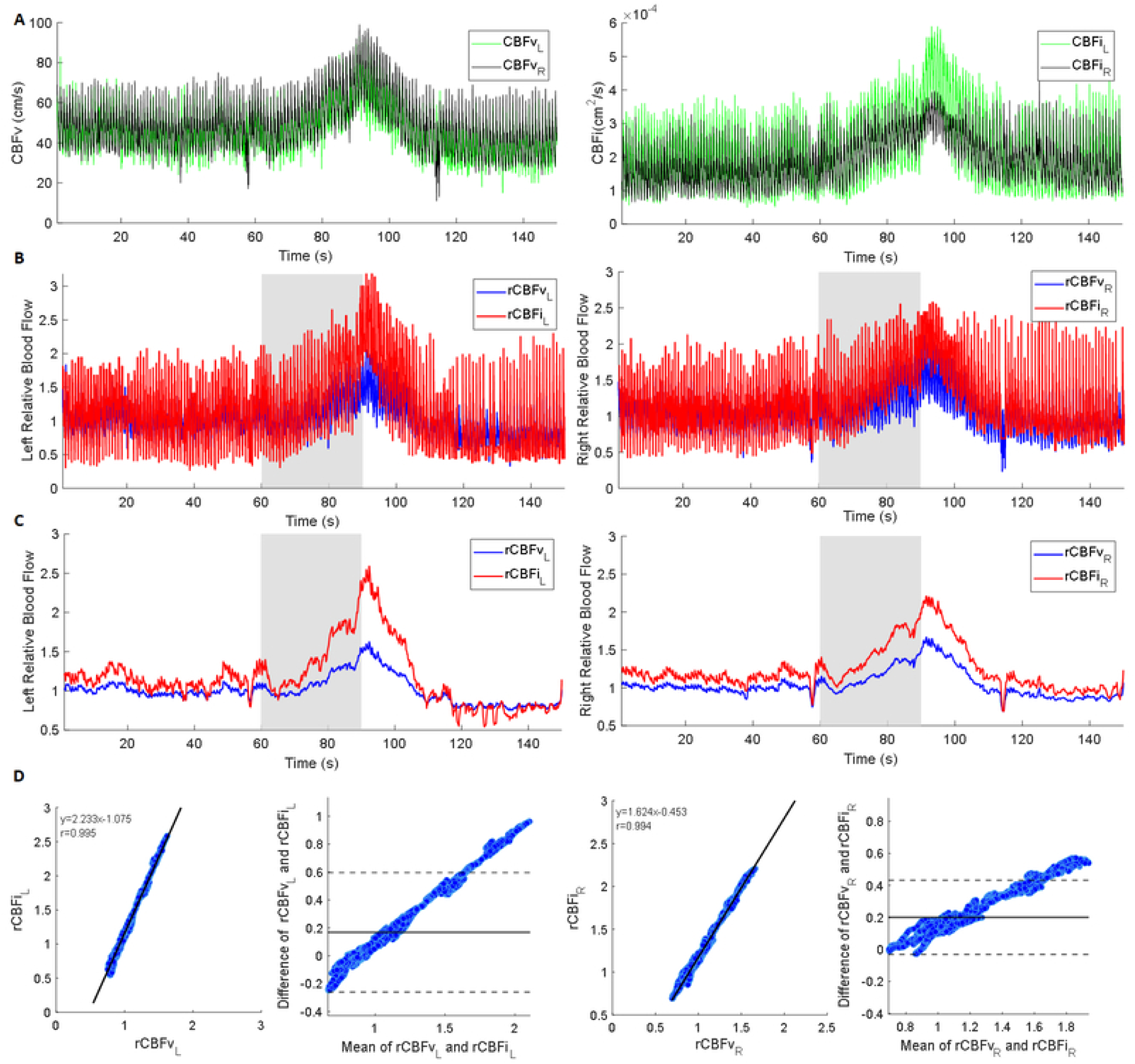
Representative single-subject analysis of TCD and DCS signals. (A) Raw bilateral middle cerebral artery CBFv signals and DCS-derived cerebral blood flow index (CBFi) signals. (B) DTW-aligned rCBFvL, rCBFvR, rCBFiL, and rCBFiR signals. (C) Smoothed rCBFvL, rCBFvR, rCBFiL, and rCBFiR signals. (D) Linear regression and Bland–Altman analyses comparing rCBFv with rCBFi for the representative participant.

### Statistical Analysis

Continuous variables were summarized as mean ± standard deviation (SD) or median (interquartile range, IQR), as appropriate. Data distributions were visualized using violin plots. All tests were two-sided, with P < 0.05 considered statistically significant. Pearson correlation quantified the association between TCD- and DCS-derived measurements. The Fisher Z transformation was applied to individual-level correlation coefficients, and the results were back-transformed to correlation coefficients for reporting. Agreement between methods was evaluated using Bland–Altman analysis, including estimation of bias and 95% limits of agreement (LoA). Linear regression analysis was performed to characterize the relationship between the two methods.

### TCD and DCS Data Processing

TCD and DCS signals were acquired synchronously and aligned using dynamic time warping. Breath-hold onset and offset were marked in the DCS trace, and raw data were stored for offline analysis. Hemodynamic parameters were extracted beat-to-beat and normalized to a 30-second baseline (starting 15 seconds after breathing stabilized), as shown in Figure 1C. Segments with low signal-to-noise ratio or motion artifacts were excluded, and remaining outliers were corrected. Relative changes from baseline were calculated for subsequent analyses.

Key hemodynamic parameters were computed to characterize cerebral perfusion responses: baseline cerebral blood flow (CBFbase, mean over 15-45 s), peak cerebral blood flow (CBFpeak, maximum within 80-100 s), relative cerebral blood flow (rCBF, calculated as the ratio of CBFpeak to CBFbase), rise time (Trise), interval from breath–hold onset at 60 s to peak perfusion), and recovery time (Trec), interval from peak to the first return–to–baseline following breath–holding. Relative cerebral blood flow was termed rCBFv for TCD and rCBFi for DCS measurements.

### Case

Figure 2 shows cerebrovascular reactivity in a representative participant (male, 33 years). As illustrated in Figure 2A and 2B, TCD-derived cerebral blood flow velocity (CBFv) and DCS-measured cerebral blood flow index (CBFi) increased in parallel during breath holding and subsequently recovered to baseline. Measurements obtained from the left and right prefrontal regions are denoted as CBFv_L_, CBFv_R_, CBFi_L_ and CBFi_R_, with left signals presented by green curves and right signals by black curves. Following dynamic time warping alignment (Figure 2C and 2D), rCBFv and rCBFi showed strong bilateral correlation (left: *r* = 0.962; right: *r* = 0.959; both *P* < 0.001), with synchronous onset, peak, and recovery. After smoothing (Figure 2E and 2F), the signals remained concordant, though rCBFi exhibited greater amplitude than rCBFv. Linear regression (Figure 2G and 2I) demonstrated strong correlations between the two methods (left: *r* = 0.995; right: *r* = 0.994; both *P* < 0.001), with slopes >1 (left: 2.215; right: 1.62). Bland–Altman analysis indicated minimal bias (left: 0.125; right: 0.200) and narrow limits of agreement (left: −0.280 to 0.528; right: −0.021 to 0.422), with no evidence of proportional bias (Figure 2H and 2J). These findings demonstrate concordant temporal dynamics between modalities. rCBFi amplitude exceeded rCBFv, while macrovascular flow and microvascular perfusion remained coupled.

### Standard Protocol Approvals, Registrations, and Patient Consents

The study protocol was approved by the Institutional Review Board of PLA General Hospital (Approval No. S2025-912-02). Written informed consent was obtained from all participants before enrollment.

### Data Availability

The data that support the findings of this study are available from the corresponding author upon reasonable request.

## Results

TCD and DCS exhibited bilaterally concordant temporal dynamics across participants (Figure 3). Linear regression revealed excellent agreement between modalities (left: r = 0.988 ± 0.009; right: r = 0.990 ± 0.008; both P < 0.001; Figure 3A, C). Bland–Altman analysis indicated minimal systematic bias and tight 95% limits of agreement (Figure 3B, D), though rCBFi displayed slight positive deviation relative to rCBFv.

**Figure 3.**
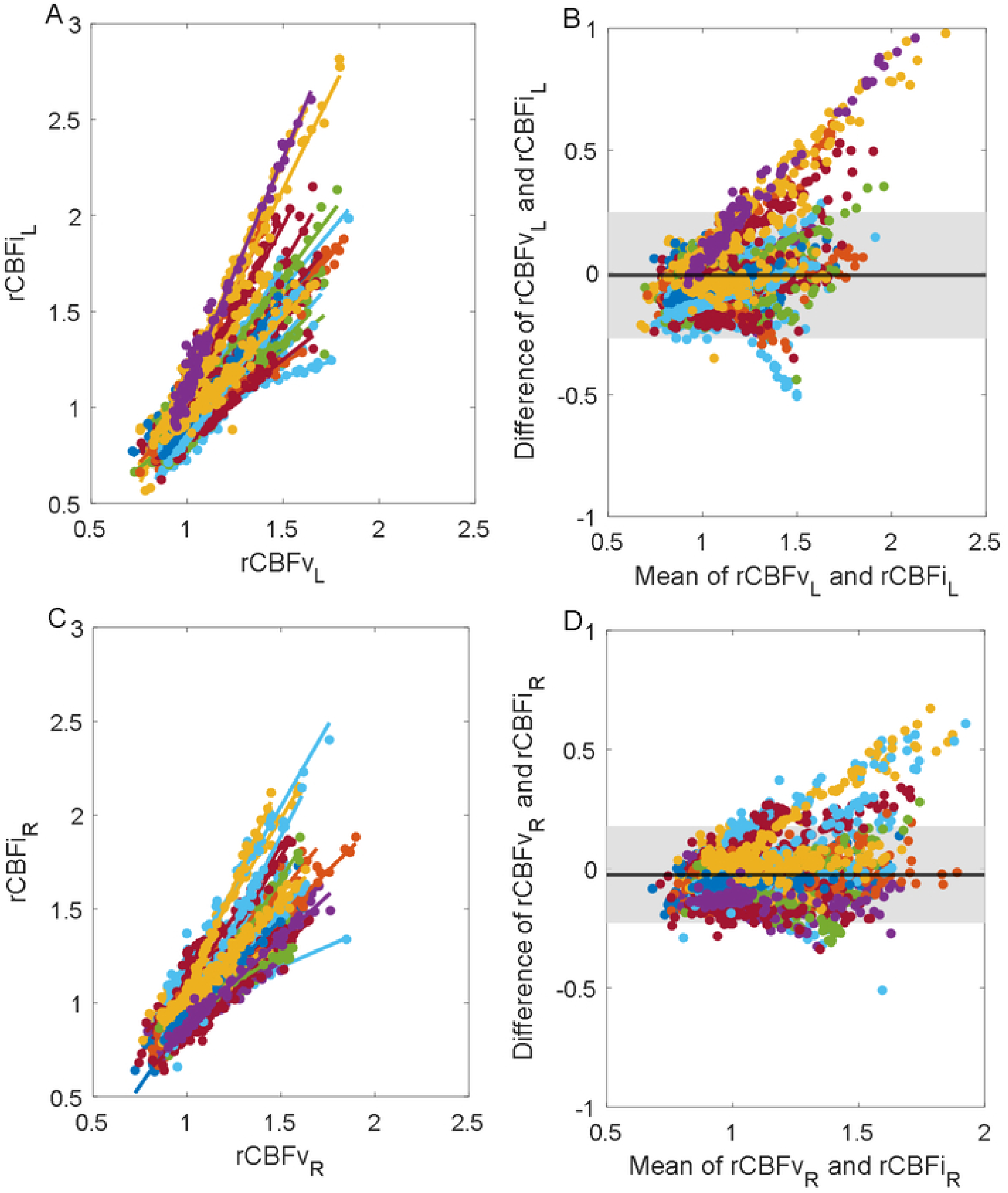
Correlation and agreement between TCD and DCS across all participants. (A) Linear regression analysis of left-sided CBFv (TCD) versus rCBF (DCS); (B) Bland–Altman analysis of left-sided data; (C) Linear regression analysis of right-sided data; (D) Bland–Altman analysis of right-sided data.

Hemodynamic parameter distributions were comparable between modalities (Figure 4). The mean values of rCBF, Trise and Trec showed no significant inter-hemispheric differences (Table 1). In contrast, DCS-derived rCBF exhibited greater variability and higher values than TCD estimates (left: 1.80 ± 0.34 vs 1.63 ± 0.15; right: 1.72 ± 0.27 vs 1.63 ± 0.14).

**Figure 4.**
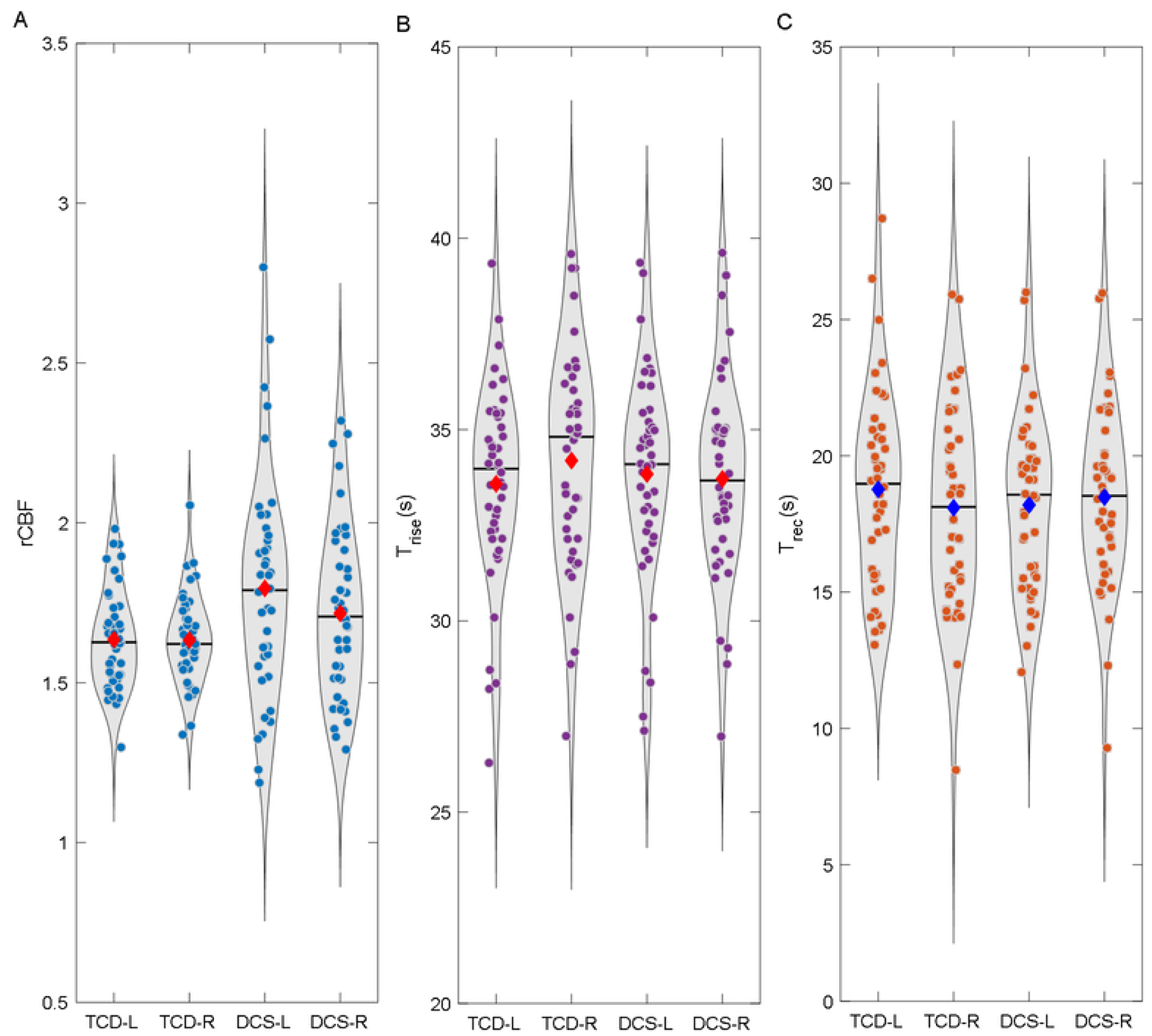
Violin plot of parameters: Distribution of blood flow parameters measured by TCD and DCS

**Table 1.** TCD- and DCS-derived cerebrovascular response parameters during breath-holding.

| Parameter | TCD-L<br>(mean±SD) | TCD-R<br>(mean±SD) | DCS-L<br>(mean±SD) | DCS-R<br>(mean±SD) |
|---|---|---|---|---|
| *rCBF* | 1.63±0.15 | 1.63±0.14 | 1.80±0.34 | 1.72±0.27 |
| *T*rise (s) | 33.58±2.60 | 34.19±2.83 | 33.84±2.71 | 33.71±2.61 |
| *T*rec (s) | 18.77±3.61 | 18.10±3.68 | 18.20±3.15 | 18.48±3.30 |

Parameter-level analyses (Figure 5) revealed strong correlations between TCD- and DCS-derived measures for Trise (left: r = 0.982; right: r = 0.883; both P < 0.001; Figure 5A, C) and Trec (left: r = 0.862; right: r = 0.890; both P < 0.001; Figure 5E, G). Bland–Altman analysis indicated minimal bias and narrow 95% limits of agreement (Figure 5B, D and 5F, H), without evidence of proportional bias.

**Figure 5.**
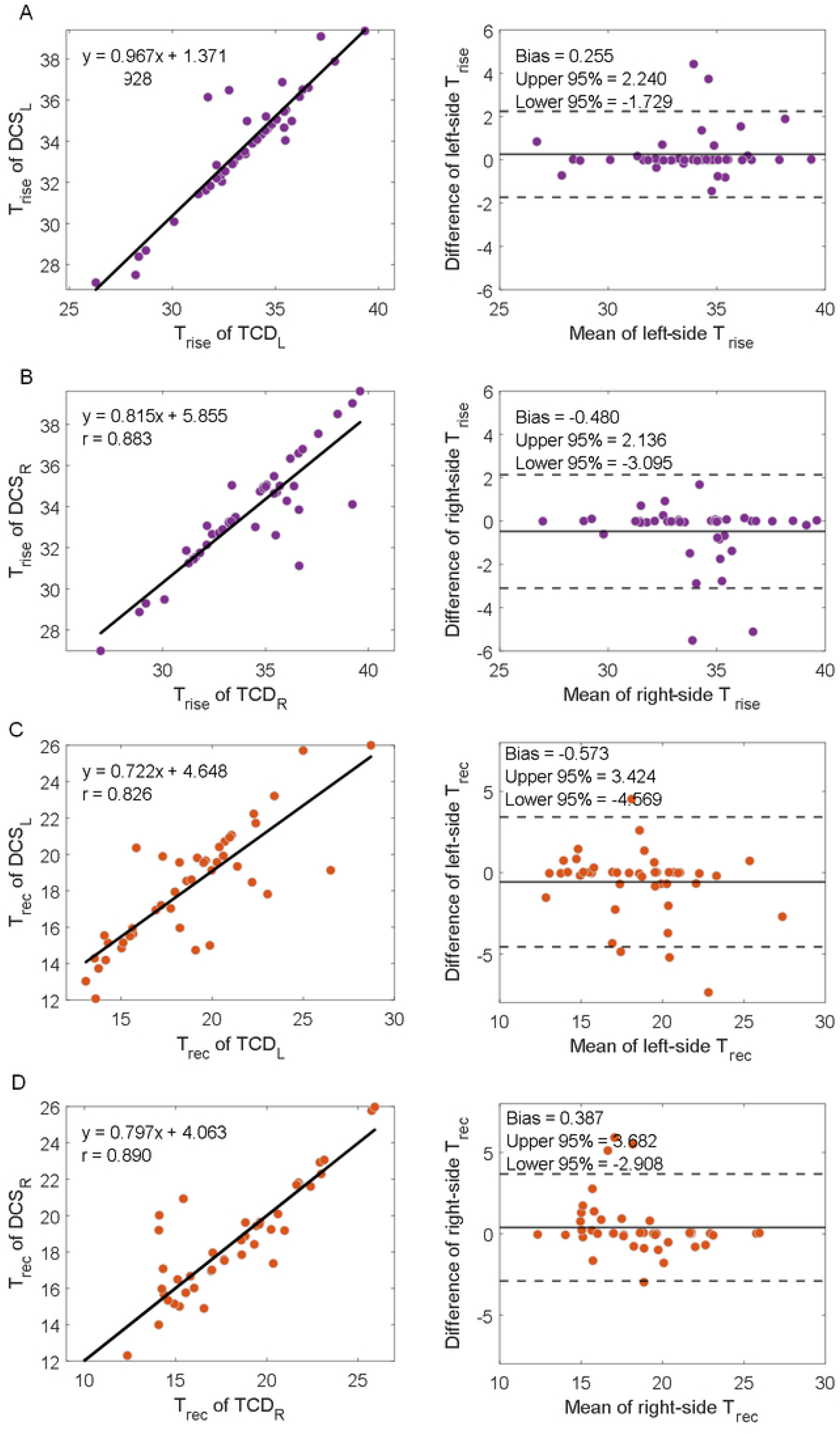
Linear regression and Bland–Altman analyses of TCD- and DCS-derived rise time (Trise) and recovery time (Trec). **(A)** Analyses of Trise for the left hemisphere. **(B)** Analyses of Trise for the right hemisphere. **(C)** Analyses of Trec for the left hemisphere. **(D)** Analyses of Trec for the right hemisphere.

Consistent with the above observations, results from the 44 participants indicate satisfactory agreement between DCS and TCD with regard to temporal dynamic characteristics, while DCS may afford superior sensitivity to microvascular dynamics.

## Discussion

In this study, simultaneous TCD-DCS monitoring showed tightly coupled temporal dynamics between TCD-derived middle cerebral artery (MCA) flow velocity and DCS-derived cortical microvascular perfusion during cerebrovascular stimulation. This concordance was evident across the full response trajectory and was supported by strong intermodality correlations, close agreement in rise and recovery times, and minimal systematic bias. These findings suggest coordinated macrovascular and microvascular responses during hypercapnic challenge, whereas differences in response magnitude indicate compartment-specific modulation of cerebrovascular reactivity.

Previous work by Zirak et al. demonstrated the feasibility of combining TCD and DCS to simultaneously assess macrovascular and microvascular cerebral hemodynamic responses during an acetazolamide challenge^15^. However, pharmacologic vasodilation differs from physiologic cerebrovascular stimulation and may be less suitable for repeated bedside assessment. In the present study, we extended this concept by integrating synchronous TCD and DCS measurements during breath-holding-induced cerebrovascular stimulation. This approach enabled noninvasive assessment of the temporal coupling between large-vessel flow velocity and cortical microvascular blood flow responses during a simple physiologic challenge.

Conventional CVR metrics typically quantify the magnitude of cerebrovascular responses by measuring stimulus-induced changes in cerebral blood flow or hemodynamic signals relative to baseline or to changes in CO₂. However, amplitude-based metrics may not fully capture the temporal characteristics of vascular responses, including response onset, time to peak, and recovery dynamics^16,17^. In contrast to approaches focused primarily on response magnitude, our study incorporated dynamic time warping analysis to characterize temporal correspondence between TCD-derived CBFv and DCS-derived CBFi responses. The strong temporal alignment observed between modalities suggests that macrovascular flow changes and microvascular perfusion responses are closely coordinated during transient hypercapnic stimulation.

We also found that DCS-derived rCBFi showed greater variability and larger relative changes than TCD-derived rCBFv. This difference likely reflects the distinct vascular compartments and spatial scales assessed by the 2 modalities rather than simple measurement disagreement. Hypercapnia induces vasodilation at multiple levels of the cerebrovascular tree, from large proximal cerebral arteries to distal arterioles and capillaries. During breath-holding-induced hypercapnia, microvascular responses may be spatially heterogeneous and may differ in magnitude from large-artery velocity changes^11,18^.Thus, one possible explanation is that DCS captures changes in red blood cell motion within the microvascular compartment, where hypercapnia-induced vasodilation and redistribution of cortical blood flow may produce larger relative signal changes than those observed in MCA flow velocity. However, the present study was not designed to identify the specific microvascular mechanisms underlying this amplitude difference.

Current approaches for evaluating cerebral hemodynamics provide important information on cerebral perfusion and vascular reserve but differ in the vascular compartments they sample and in their ability to capture dynamic responses over time ^19–21^. By capturing both response magnitude and temporal coordination, simultaneous TCD-DCS monitoring may provide a more comprehensive assessment of cerebrovascular function than either modality alone.Our findings suggest that assessment of cerebrovascular regulation may require characterization of the temporal coordination of responses across vascular compartments. Potential clinical applications include continuous cerebrovascular monitoring in neurocritical care, evaluation of patients with inadequate TCD acoustic windows, dynamic hemodynamic assessment in acute ischemic stroke, characterization of microvascular dysfunction in cerebral small vessel disease and vascular cognitive impairment, and longitudinal bedside monitoring of cerebral microcirculation. Whether these measures provide incremental value for risk stratification or outcome prediction requires evaluation in prospective clinical cohorts. Several limitations should be considered. First, this study was conducted in a relatively small cohort of healthy young adults; therefore, the findings require validation in larger and more diverse populations, particularly in patients with cerebrovascular disease. Second, TCD and DCS sample different vascular compartments and spatial scales. TCD measures MCA flow velocity rather than absolute cerebral blood flow, whereas DCS reflects regional cortical microvascular perfusion within a limited sampling volume. Accordingly, differences in response magnitude between rCBFv and rCBFi should be interpreted cautiously. Third, preprocessing steps, including signal smoothing and normalization, may influence estimates of response amplitude and temporal parameters. Fourth, these findings were obtained during a breath-holding hypercapnic stimulus and may not generalize to other CVR paradigms, such as acetazolamide challenge, controlled CO₂ inhalation, breath-holding protocols of different duration, or task-based neurovascular activation. Finally, although the observed temporal concordance supports macrovascular-microvascular coupling, this study cannot establish the underlying regulatory mechanisms. Future studies incorporating larger clinical cohorts, complementary perfusion imaging, standardized stimulation protocols, and longitudinal outcome assessment are needed to confirm the physiologic and clinical relevance of these dynamic coupling metrics.

## Conclusion

In conclusion, simultaneous TCD-DCS monitoring provides a feasible multimodal approach for assessing both large-vessel flow velocity and cortical microvascular perfusion during hypercapnic stimulation. The closely aligned temporal profiles of TCD-derived rCBFv and DCS-derived rCBFi support macrovascular–microvascular coupling, whereas differences in response magnitude suggest compartment-specific vascular regulation. These findings provide a physiologic basis for future studies evaluating TCD-DCS monitoring as a bedside tool for dynamic assessment of cerebrovascular reactivity and microvascular function.

## Disclosure

The authors have no conflicts of interest to disclose.

## Author Contributions

Zhe Ji: drafting of the manuscript, including medical writing for content; analysis or interpretation of data. HuiSu: critical revision of the manuscript for intellectual content; analysis or interpretation of data. Yiwen Wang: major role in the acquisition of DCS data; analysis or interpretation of data. Yiqin Han: major role in the acquisition of TCD data; data verification. ZheLi: analysis or interpretation of data; data curation; critical revision of the manuscript for intellectual content. Xiangyu Cao: study concept or design; study supervision; overall coordination of the study; critical revision of the manuscript for intellectual content.

